# Putative Causal Effects of Amyloid-β on GFAP: a Mendelian Randomization Study

**DOI:** 10.64898/2026.11.25.26368912

**Authors:** Ron Danziger, Pia Kivisäkk, Ling Teng, Zheng Yin, Jingyun Yang, Lei Yu, Jijing Wang, David H. Adamowicz, Brahyan J. Galindo-Mendez, Dylan V. Neel, Wai-Ying Wendy Yau, Dennis J. Selkoe, Julie A. Schneider, David A. Bennett, Philip L. De Jager, Rachel F. Buckley, Keith A. Johnson, Reisa A. Sperling, Steven E. Arnold, Jasmeer P. Chhatwal, Hyun-Sik Yang

**Affiliations:** Department of Neurology, Mass General Brigham, Boston, MA, USA; Harvard Medical School, Boston, MA, USA; Broad Institute of Harvard and Massachusetts Institute of Technology, Cambridge, MA, USA; Rush Alzheimer’s Disease Center, Rush University Medical Center, Chicago, IL, USA; Department of Neurological Sciences, Rush University Medical Center, Chicago, IL, USA; Department of Psychiatry, Mass General Brigham, Boston, MA, USA; Department of Pathology, Rush University Medical Center, Chicago, IL, USA; Department of Neurology, Columbia University Irving Medical Center, New York, NY, USA; Department of Radiology, Mass General Brigham, Boston, MA, USA

**Author notes:** Correspondence to: Hyun-Sik Yang, MD, Center for Alzheimer Research and Treatment, Department of Neurology, Brigham and Women’s Hospital, 60 Fenwood Road, Boston, MA 02115, USA.

## Abstract

**INTRODUCTION:** Glial fibrillary acidic protein (GFAP) is a marker of astrocytic activation that correlates with cerebral Alzheimer disease (AD) pathology. The causal relationship between GFAP elevation and amyloid-β (Aβ) remains unclear. We examined whether Aβ drives GFAP elevation and whether GFAP is independently associated with AD progression.

**METHODS:** In the Harvard Aging Brain Study (HABS; n=329 cognitively unimpaired participants) and the Religious Orders Study and Rush Memory and Aging Project (ROSMAP; n=835 deceased participants spanning cognitively unimpaired to dementia), we performed one-sample Mendelian randomization. *APOE* ε4 served as the instrumental variable to estimate the causal effect of Aβ (PET in HABS; immunohistochemistry in ROSMAP) on GFAP (plasma in HABS; brain mRNA expression in ROSMAP). Linear regression and linear mixed-effects models assessed GFAP associations with tau, hippocampal atrophy, and cognitive decline.

**RESULTS:** In both cohorts, Mendelian randomization supported a causal effect of Aβ on GFAP (HABS β=0.37; 95% CI, 0.11 to 0.63; *P*=6.1×10⁻³; ROSMAP β=0.35; 95% CI, 0.12 to 0.59; *P*=3.3×10⁻³). In HABS, GFAP associations with tau and cognition were no longer significant after adjusting for Aβ, and GFAP was not associated with hippocampal atrophy. In ROSMAP, GFAP remained associated with tau and cognition after Aβ adjustment, but not with cognition after additional adjustment for tau.

**DISCUSSION:** Brain GFAP gene expression and plasma GFAP elevation may be downstream consequences of Aβ accumulation. GFAP showed little independent association with cognition once core AD pathology was accounted for. These findings refine the interpretation of elevated plasma GFAP in AD.

## 1.0 Introduction

For much of the past two decades, detection of Alzheimer disease (AD) pathology *in vivo* required costly or invasive testing such as cerebrospinal fluid (CSF) analysis or amyloid positron emission tomography (PET).^1,2^ Sensitive plasma immunoassays now allow AD biomarkers to be quantified from a routine blood draw, even in the preclinical phase.^3,4^ The 2024 Alzheimer’s Association criteria incorporated blood-based biomarkers of amyloid and tau, such as plasma phosphorylated tau 217 (p-tau217),^5,6^ into AD diagnosis and staging.^7^ The criteria also designate CSF and plasma glial fibrillary acidic protein (GFAP) as a biomarker of inflammation (“I”), an arm of AD pathology increasingly recognized as an active contributor to disease progression.^7,8^ Plasma GFAP is widely used in clinical studies, shows a robust association with cerebral amyloid-β (Aβ), and is thought to reflect reactive astrogliosis surrounding amyloid plaques.^9–12^ However, whether elevated GFAP reflects core AD pathophysiology or merely correlates with it remains poorly defined. Plasma GFAP has been associated with cognitive decline and dementia,^13–16^ but it is unclear whether GFAP adds diagnostic or prognostic value over clinically established biomarkers.

GFAP is a nonspecific marker of astrocytic activation and injury and is elevated across many neurological conditions.^17^ With ubiquitin C-terminal hydrolase L1 (UCH-L1), GFAP is FDA-approved for detecting intracranial injury after traumatic brain injury and is elevated in progranulin (*GRN*) mutation-related frontotemporal dementia.^18,19^ Thus, what an elevated GFAP level captures in the specific context of human AD requires deeper study. For example, Aβ may drive astrocytic activation and GFAP expression, or conversely, reactive astrocytes may contribute to Aβ accumulation.^20–22^ Anti-amyloid therapies lower plasma GFAP in proportion to Aβ plaque clearance, suggesting a causal link, but the causal relationship between Aβ and GFAP in the natural history of AD remains to be clarified.^23^ While this relationship is difficult to infer from simple statistical correlations, Mendelian randomization (MR) can be used to estimate a putative causal effect under specific assumptions, using genetic variants as instrumental variables to reduce confounding and reverse causation.^24–27^ APOE ε4, the strongest common genetic risk factor for late-onset AD, is robustly associated with earlier and greater cerebral Aβ deposition, which makes it a suitable genetic instrument for Aβ burden.^28^

Here, we applied MR across two independent cohorts, integrating *in vivo* plasma and imaging biomarkers with postmortem histopathological and transcriptomic data, to estimate the causal effect of Aβ on GFAP using *APOE* ε4 as a single instrumental variable. We then examined GFAP’s associations with tau pathology, neurodegeneration, and cognitive decline, before and after accounting for Aβ burden.

## 2.0 Methods

### 2.1 Participants

We included participants from two cohorts, the Harvard Aging Brain Study (HABS) and the Religious Orders Study and Rush Memory and Aging Project (ROSMAP). HABS is a prospective cohort study of older adults aged 50–90 years and cognitively unimpaired (CU) at baseline, conducted in Boston since 2010, with annual cognitive testing and blood sample collection and neuroimaging every 3 years. Inclusion and exclusion criteria have been previously described.^29,30^ Only participants with available plasma GFAP, ^11^C-Pittsburgh compound B (PiB) PET, and *APOE* genotype data at baseline were included. HABS was approved by the Mass General Brigham Institutional Review Board (IRB), and all participants provided written informed consent.

ROSMAP is a combined dataset from two longitudinal cohort studies conducted at the Rush Alzheimer’s Disease Center: the Religious Orders Study (ROS, launched 1994), which enrolls older Catholic clergy across the United States, and the Rush Memory and Aging Project (MAP, launched 1997), which recruits older adults from retirement communities and senior housing throughout the greater Chicago metropolitan area and northeast Illinois. Both studies enroll participants without known dementia at baseline and follow them annually until death with longitudinal clinical and cognitive evaluations. Upon death, brain tissue is collected at autopsy for molecular analysis. Further details of ROSMAP have been previously described.^31,32^ Only participants with postmortem RNA-sequencing (RNA-seq) data from the dorsolateral prefrontal cortex (DLPFC), quantitative Aβ immunohistochemistry (IHC), and *APOE* genotype data were included. ROS and MAP were approved by a Rush University Medical Center IRB, and all participants provided written informed and repository consents and signed an Anatomical Gift Act for brain donation.^31^

### 2.2 Procedures: Harvard Aging Brain Study (HABS)

Plasma GFAP was measured using the Meso Scale Discovery (MSD) Neurology 3-PLEX Array (MSDN3). The assay was run in duplicate, and we included measures with a coefficient of variation (CV) <20%.^33,34^ *APOE* genotype was determined through direct genotyping of *APOE* codons 112 and 158.^29,30^

In vivo Aβ plaque burden was measured with PiB PET and in vivo fibrillar tau burden was measured using ^18^F-flortaucipir (FTP) PET (tau PET) at the Massachusetts General Hospital on an ECAT EXACT HR+ scanner (Siemens). PET scans were performed at study years 1, 4, 6, 9, and 12.^35^ As FTP-PET became available mid-study, participants in HABS underwent their first tau PET at 2.4±1.6 years after the baseline visit. Regions of interest (ROIs) were defined using FreeSurfer (version 6.0). Aβ burden was measured as a distribution volume ratio (DVR) without partial volume correction (PVC) from a composite across frontal, lateral temporal, parietal, and retrosplenial regions (FLR). Tau burden was measured with a standardized uptake value ratio (SUVR) with PVC within the inferior temporal cortex (ITC), where early neocortical tau accumulation is observed in preclinical AD.^36,37^ Both PiB PET and tau PET used cerebellar gray matter as the reference region.

Bilateral hippocampal volume (HV) was measured with structural magnetic resonance imaging (MRI) on a 3T Magnetom Trio TIM scanner (Siemens) and was adjusted for total intracranial volume, as previously described.^37^

The Preclinical Alzheimer Cognitive Composite-5 (PACC5) was administered annually for longitudinal cognitive assessment.^38^ PACC5 is a composite that includes five cognitive tests (Mini-Mental State Examination, Logical Memory delayed recall, Free and Cued Selective Reminding Test, Digit Symbol Substitution, and Category Fluency) to detect early Aβ-related cognitive decline.^39–43^

### 2.3 Procedures: ROSMAP

Codons 112 and 158 from *APOE* were sequenced to derive *APOE* haplotypes (ε2/ε3/ε4).^44^ GFAP mRNA levels were derived from bulk RNA sequencing of frozen brain tissue from the DLPFC.^45^ Raw FASTQ reads were aligned to the GRCh38 reference genome (GENCODE v24) and quantified with STAR, yielding per-sample count matrices and coordinate-sorted, indexed BAM files. Alignment and expression-level quality were evaluated with Picard. Samples were removed if they had an RNA integrity number (RIN) <4, low mean sequencing quality, or a postmortem interval (PMI) ≥24 hours, as were samples with duplicate participant identifiers. Features were restricted to genes exceeding 1 count per million in at least 50% of samples, and counts were normalized for gene length and GC content by conditional quantile normalization (CQN). No outliers were identified by principal component analysis or hierarchical clustering. Expression abundances and observation-level precision weights were then estimated with the voom function in the limma package^46^ and residualized against batch, RIN, PMI, sex, age at death, and sequencing-quality metrics (the proportion of aligned reads and the proportions of intergenic, intronic, and coding bases). The resulting residual expression matrix served as the input for all downstream analyses. Aβ was quantified via IHC as the mean percent area occupied by Aβ in the midfrontal cortex (the region where GFAP gene expression was quantified). Tau was quantified using anti-phospho-tau (AT8) antibody in the same region and expressed as mean cortical density (per mm^2^).^32^ In addition, for sensitivity analyses, we assessed neuritic plaques, diffuse plaques, and neurofibrillary tangles (NFTs), quantified using a modified Bielschowsky silver stain. All quantitative neuropathology variables were square-root-transformed to reduce skew.^32^

Longitudinal cognitive change was captured using a “global cognitive slope” of average z-standardized test scores across 19 cognitive tests administered annually. Individual rates of cognitive change were estimated as person-specific random slopes from a linear mixed model of global cognition over time, controlling for age at baseline, sex, education, and whether a telephone evaluation was used.^47^ Because this person-specific random slope requires longitudinal cognitive assessment, it could not be estimated for participants with insufficient follow-up. These individuals were excluded from cognition analyses. Final consensus clinical diagnosis proximate to death was used to classify participants as CU, mild cognitive impairment (MCI), or dementia.^31,32^

### 2.4 Statistical Analysis

Analyses were performed in R (version 4.5.1). Participants with missing data were excluded. Variables were z-standardized. All models were adjusted for age and sex, and models with cognitive outcomes were additionally adjusted for years of education, either through inclusion as covariates or residualization (GFAP mRNA and global cognitive slope in ROSMAP). MR was performed in each cohort using *APOE* ε4 carrier status as the instrumental variable (IV), Aβ as the exposure, and GFAP as the outcome. In HABS, the exposure was the baseline PiB PET cortical composite, and the outcome was baseline plasma GFAP; in ROSMAP, the exposure was postmortem IHC-quantified Aβ burden, and the outcome was GFAP mRNA expression. Estimated causal effects were obtained by two-stage least squares (TSLS) using the “ivmodel” package. The goal of primary MR analyses was to derive the cohort-specific causal estimate of Aβ on GFAP.

MR assumptions (relevance, independence, and exclusion restriction) were evaluated in accordance with STROBE-MR guidelines.^25,26^ Relevance was assessed by regressing Aβ on *APOE* ε4, using an *F*>10 threshold to assess instrument strength.^48^ Independence is supported by the germline autosomal inheritance of *APOE* ε4, fixed at conception and unaffected by postnatal factors.^28^ To detect gross violations of the exclusion restriction, the association between *APOE* ε4 and GFAP was assessed after adjusting for Aβ; we reasoned that the *APOE* ε4-GFAP association would remain significant if there were important Aβ-independent pathways linking *APOE* ε4 to elevated GFAP.

To assess the robustness of the primary ROSMAP estimate, we performed sensitivity and replication analyses. To rule out results driven by population stratification, the ROSMAP MR was repeated among participants of European ancestry, with adjustment for genetic ancestry principal components. To assess whether the causal estimate differed by disease stage, the ROSMAP MR was repeated within each clinical diagnosis (CU, MCI, and dementia). To confirm the core findings were not specific to the immunohistochemical measures, we replicated the primary analyses using silver-stain neuropathology, with midfrontal neuritic plaque burden (as the measure of Aβ), defined as the exposure, and GFAP mRNA expression defined as the outcome. To determine whether the causal effect of Aβ on GFAP is also present in the absence of peri-plaque neuritic response, we repeated the MR using midfrontal diffuse plaque burden, a non-fibrillar amyloid species that is bound only weakly by amyloid PET ligands.^49^ In this case, diffuse plaque burden was defined as the exposure and GFAP mRNA expression as the outcome. To compare the two plaque types directly, GFAP mRNA expression was regressed on neuritic and diffuse plaque burden in the same model.

Secondary analyses examined the association of GFAP with other indicators of AD pathophysiology. In HABS, participants with repeated tau PET, MRI, or PACC5 assessments were analyzed using linear mixed-effects models (“lme4” and “lmerTest” packages; with random intercepts for tau PET and MRI and random intercepts and slopes for PACC5) with a predictor-by-time interaction, testing whether GFAP was associated with longitudinal change in tau PET (ITC) signal, hippocampal volume, and PACC5. In ROSMAP, the relationship of GFAP mRNA expression with quantitative tau IHC and global cognitive slope was assessed by linear regression. We then examined whether the observed association between GFAP and AD traits was independent of Aβ and tau by including Aβ (tau and cognition outcomes) and tau (cognition outcome) as covariates in the model. For these adjusted models, we report the partial R² for GFAP, the proportion of residual variance in the outcome explained by GFAP after accounting for the other covariates. Next, to test whether the GFAP-tau and GFAP-cognition relationships differed by disease stage in ROSMAP, these analyses were repeated, stratified by clinical diagnosis (CU, MCI, and dementia). Finally, the ROSMAP GFAP-tau and GFAP-cognition analyses were repeated with midfrontal NFT burden (silver stain), each before and after adjustment for neuritic plaque burden (silver stain). The cognition analysis was additionally adjusted for NFT burden.

## 3.0 Results

### 3.1 Participant characteristics

We included 329 HABS participants who had baseline PiB PET and plasma GFAP (mean age 71.5±8.0 years; 62% female; 28% *APOE* ε4 carriers) and 835 ROSMAP participants with complete autopsy neuropathology and DLPFC RNA-seq data (mean age at enrollment 80.9±7.0 years; mean age at death 89.1±6.6 years; 64% female; 26% *APOE* ε4 carriers) (**Table 1**, **Figure 1**).

**Figure 1.**
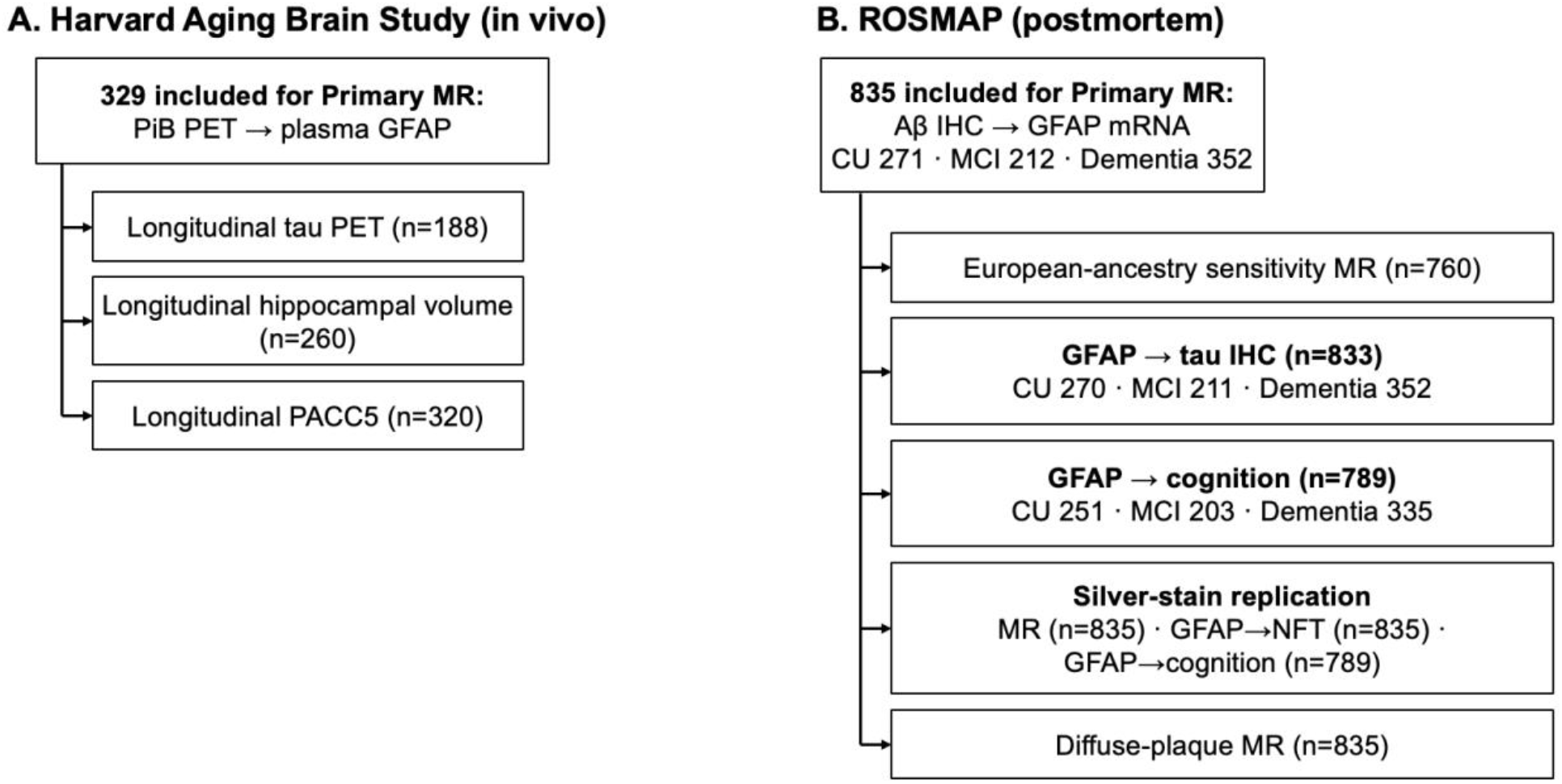
Participant selection. Analytic samples from [A] the Harvard Aging Brain Study (HABS) and [B] the Religious Orders Study and Rush Memory and Aging Project (ROSMAP). The top box in each panel shows the primary Mendelian randomization sample. Boxes below show the secondary analyses and their sample sizes. For ROSMAP, the numbers of cognitively unimpaired (CU), mild cognitive impairment (MCI), and dementia participants are shown for the stage-stratified analyses. **Abbreviations**: Aβ, amyloid-β; CU, cognitively unimpaired; GFAP, glial fibrillary acidic protein; IHC, immunohistochemistry; MCI, mild cognitive impairment; MR, Mendelian randomization; NFT, neurofibrillary tangle; PACC5, Preclinical Alzheimer Cognitive Composite-5; PET, positron emission tomography; PiB, Pittsburgh compound B.

**Table 1.** Participant characteristics.

| **Characteristic** | **HABS (*n*=329)** | **ROSMAP (*n*=835)** |
|---|---|---|
| Age, mean ± SD | 71.5±8.0 | 80.9±7.0 (enrollment)<br>89.1±6.6 (death) |
| Female, *n* (%) | 203 (62) | 538 (64) |
| Race, *n* (%) | White, 254 (77)<br>Black, 49 (15)<br>Asian, 7 (2)<br>Other/mixed, 5 (2)<br>Not reported, 14 (4) | White, 822 (98)<br>Black, 12 (1)<br>Other, 1 (<1) |
| Years of education, mean ± SD | 15.8±2.9 | 16.4±3.6 |
| *APOE* ε4 carrier, *n* (%) | 91 (28) | 217 (26) |
| MMSE, median (IQR) | 29 (29–30) | 25 (16–28) |
| AD pathology, *n* (%) | PiB PET+ 77 (23)<br>PiB PET− 252 (77) | CERAD moderate/frequent 530 (63)<br>CERAD none/sparse 305 (37) |
| Clinical diagnosis, *n* (%) | CU, 329 (100) | CU, 271 (32)<br>MCI, 212 (25)<br>Dementia, 352 (42) |

**Abbreviations**: AD, Alzheimer disease; APOE, apolipoprotein E; CERAD, Consortium to Establish a Registry for Alzheimer Disease; CU, cognitively unimpaired; HABS, Harvard Aging Brain Study; IQR, interquartile range; MCI, mild cognitive impairment; MMSE, Mini-Mental State Examination; PET, positron emission tomography; PiB, Pittsburgh compound B; ROSMAP, Religious Orders Study and Rush Memory and Aging Project; SD, standard deviation.

### 3.2 Putative causal effect of Aβ on GFAP across two distinct cohorts

In HABS, *APOE* ε4 was robustly associated with PiB PET (β=0.76; 95% CI, 0.54 to 0.98; *P*=7.3×10⁻¹¹; *F*=45.4). *APOE* ε4 was also associated with plasma GFAP (β=0.31; 95% CI, 0.10 to 0.52; *P*=3.7×10⁻³), but this association was no longer significant after adjustment for Aβ (*P*=0.35). TSLS analysis estimated the causal effect of Aβ on plasma GFAP (β=0.37; 95% CI, 0.11 to 0.63; *P*=6.1×10⁻³) (**Figure 2, A and B**).

**Figure 2.**
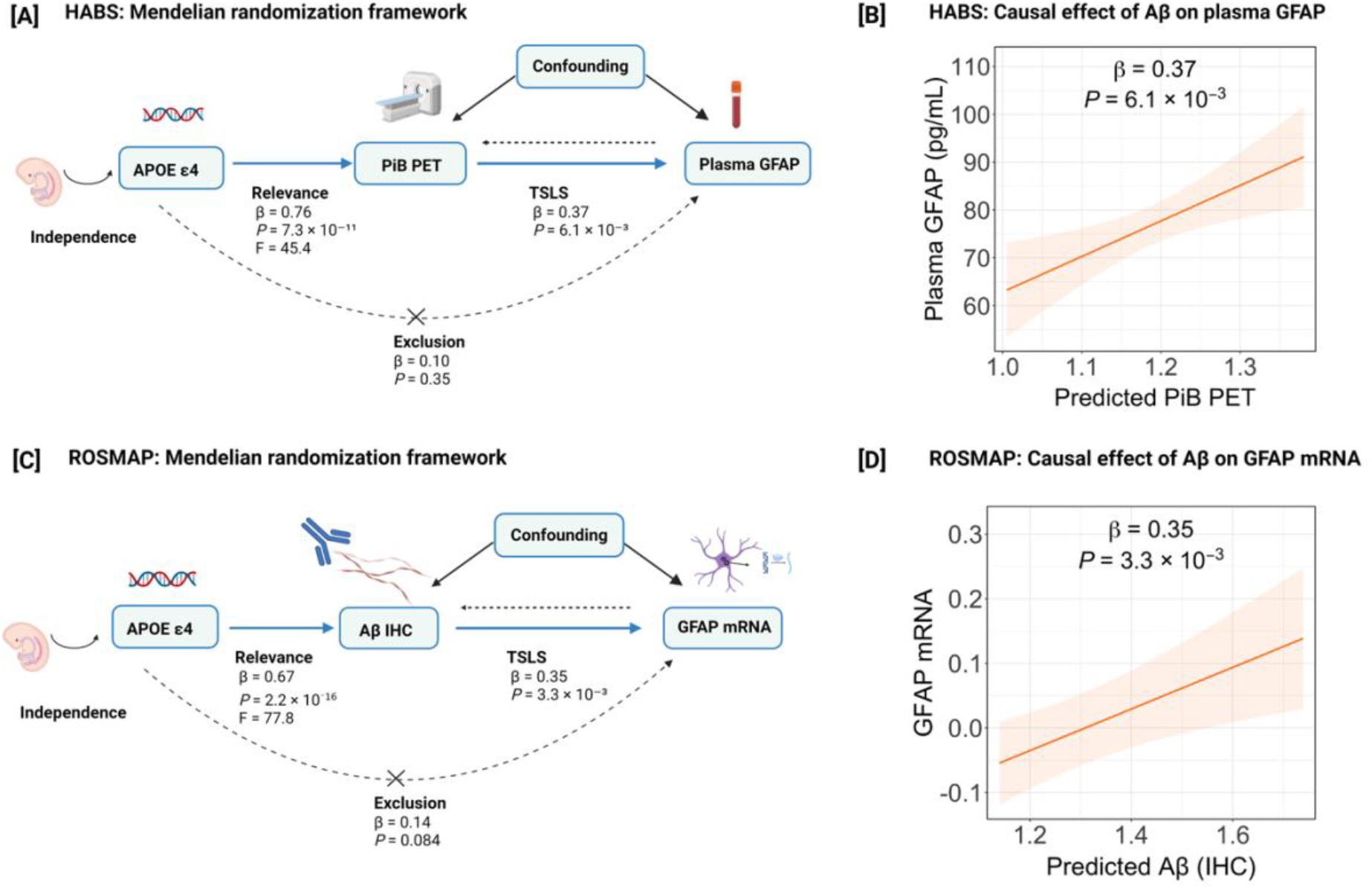
Mendelian randomization of amyloid-β on GFAP. APOE ε4 carrier status served as a genetic instrumental variable for the causal effect of amyloid-β (Aβ) on GFAP. HABS models were adjusted for age and sex. In ROSMAP models, GFAP mRNA expression was pre-residualized as described in Methods. In the diagrams, solid blue arrows denote the instrument-exposure association (Relevance) and the two-stage least squares (TSLS) causal estimate. The backward dashed arrow denotes reverse causation (excluded by design); the crossed dashed arrow (X) is the exclusion-restriction path, for which the values shown are the APOE ε4-GFAP association after adjustment for Aβ. The Confounding box denotes unmeasured confounders of the Aβ-GFAP relationship, which the instrument is assumed to be independent of. Independence is supported by APOE ε4 being germline and fixed at conception. [A] Instrumental-variable diagram for HABS: PiB PET cortical composite is the exposure and plasma GFAP the outcome. [B] Fitted second-stage relationship between genetically predicted Aβ and plasma GFAP (HABS). [C] Instrumental-variable diagram for ROSMAP: quantitative Aβ immunohistochemistry (IHC) is the exposure and dorsolateral prefrontal cortex GFAP mRNA the outcome. [D] Fitted second-stage relationship between genetically predicted Aβ and GFAP mRNA (ROSMAP). Shaded orange bands are 95% confidence intervals of the fitted second-stage regression; all β are standardized. **Abbreviations**: Aβ, amyloid-β; APOE, apolipoprotein E; GFAP, glial fibrillary acidic protein; HABS, Harvard Aging Brain Study; IHC, immunohistochemistry; mRNA, messenger RNA; PET, positron emission tomography; PiB, Pittsburgh compound B; ROSMAP, Religious Orders Study and Rush Memory and Aging Project; TSLS, two-stage least squares. Created in BioRender. Danziger, R. (2026) https://BioRender.com/6m7hfmu

In ROSMAP, *APOE* ε4 was robustly associated with Aβ burden (β=0.67; 95% CI, 0.52 to 0.81; *P*=2.2×10⁻¹⁶; *F*=77.8). *APOE* ε4 was also associated with GFAP mRNA expression (β=0.23; 95% CI, 0.080 to 0.39; *P*=2.9×10⁻³), but this association was no longer significant after adjustment for Aβ (*P*=0.084). TSLS analysis estimated the causal effect of Aβ on GFAP mRNA expression (β=0.35; 95% CI, 0.12 to 0.59; *P*=3.3×10⁻³) (**Figure 2, C and D**). A sensitivity analysis restricted to participants of European ancestry, adjusting for genetic ancestry principal components, yielded similar results (n=760, β=0.34; 95% CI, 0.096 to 0.59; *P*=6.5×10⁻³; *F*=68.9). In an MR stratified by clinical diagnosis, the causal estimate of Aβ on GFAP mRNA expression was similar across stages (CU, β=0.39; MCI, β=0.28; dementia, β=0.31) and consistent with the overall estimate, although none reached significance within an individual stage (**Supplementary Figure 1**).

### 3.3 Association of plasma GFAP with longitudinal tau, hippocampal volume, and cognition in HABS

Baseline plasma GFAP was associated with longitudinal tau PET among participants with repeated scans (*n*=188, β=0.021; 95% CI, 0.0040 to 0.039; *P*=0.017) (**Figure 3A**); however, the association became nonsignificant after adjustment for Aβ (*P*=0.46) (**Figure 3B**). Baseline plasma GFAP was not associated with longitudinal bilateral hippocampal volume, before (*n*=260, *P*=0.52) (**Figure 3C**) or after Aβ adjustment (*P*=0.30) (**Figure 3D**). Baseline plasma GFAP was associated with faster PACC5 decline (*n*=320, β=−0.030; 95% CI, −0.048 to −0.012; *P*=1.5×10⁻³) (**Figure 3E),** but this association was no longer significant after adjustment for Aβ (*P*=0.85) (**Figure 3F**).

**Figure 3.**
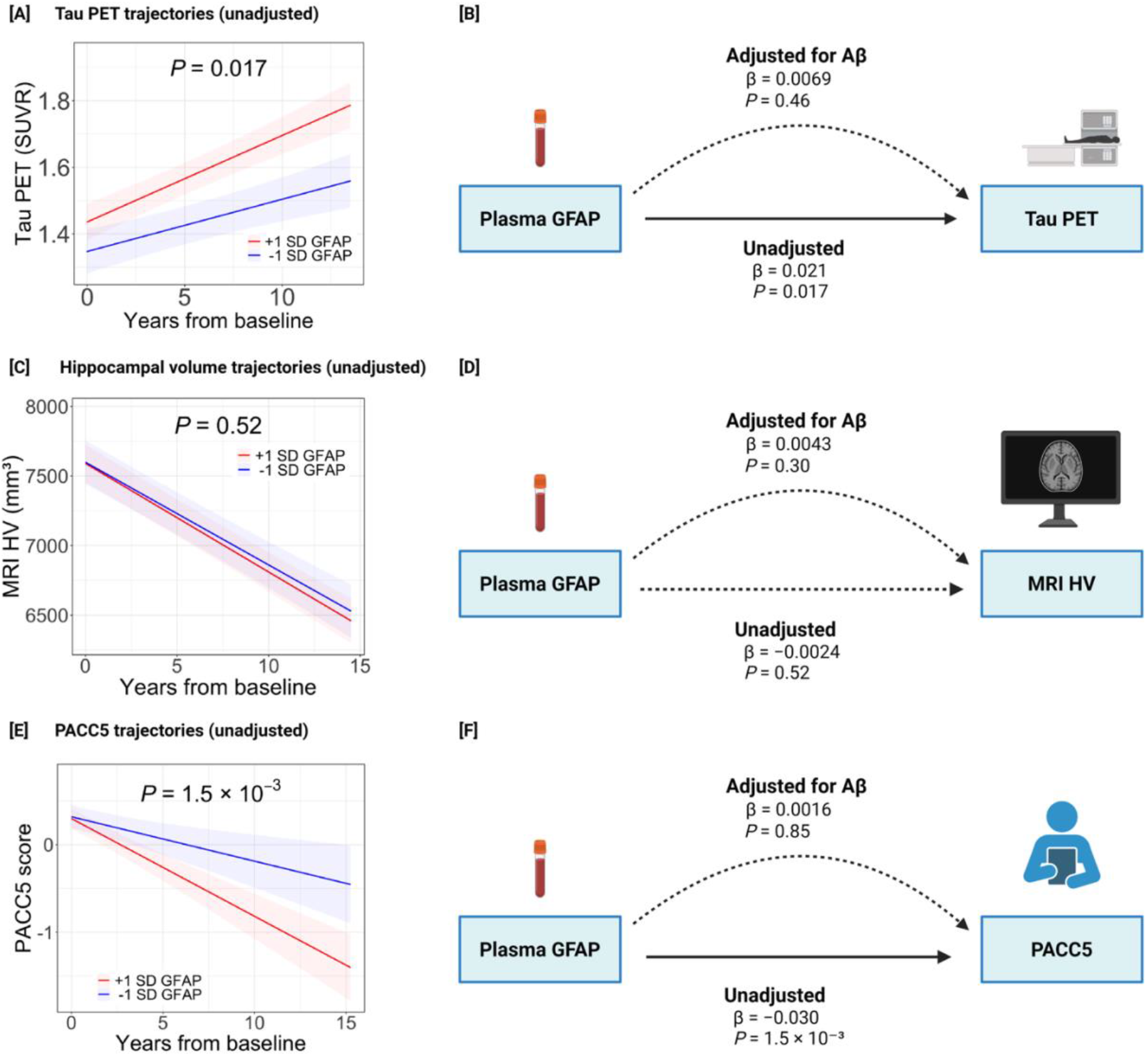
Association of baseline plasma GFAP with longitudinal tau, neurodegeneration, and cognition in cognitively unimpaired adults (HABS). Linear mixed-effects models tested whether baseline plasma GFAP was associated with longitudinal change in each outcome, before and after adjustment for amyloid-β (Aβ); all models were adjusted for age and sex (and years of education for PACC5). In the trajectory plots, lines show model-predicted change at +1 SD (red) and −1 SD (blue) plasma GFAP, shaded bands are 95% confidence intervals, and the annotated P value is for the GFAP × time interaction (unadjusted for Aβ). In the diagrams, straight arrows denote the unadjusted association and curved arrows the Aβ-adjusted association; solid arrows indicate statistical significance (P<0.05) and dashed arrows nonsignificance. The β and P values shown are for the standardized GFAP × time interaction. [A] Tau PET (inferior temporal cortex) trajectories by GFAP level. [B] Plasma GFAP and longitudinal tau PET, before and after Aβ adjustment. [C] Hippocampal volume trajectories by GFAP level. [D] Plasma GFAP and longitudinal hippocampal volume, before and after Aβ adjustment. [E] PACC5 trajectories by GFAP level. [F] Plasma GFAP and longitudinal PACC5, before and after Aβ adjustment. **Abbreviations**: Aβ, amyloid-β; GFAP, glial fibrillary acidic protein; HABS, Harvard Aging Brain Study; HV, hippocampal volume; MRI, magnetic resonance imaging; PACC5, Preclinical Alzheimer Cognitive Composite-5; PET, positron emission tomography; SD, standard deviation; SUVR, standardized uptake value ratio. Created in BioRender. Danziger, R. (2026) https://BioRender.com/0g51fxn

### 3.4 Association of GFAP mRNA expression with tau pathology and cognition in ROSMAP

GFAP mRNA expression was associated with quantitative tau IHC (*n*=833, β=0.15; 95% CI, 0.080 to 0.21; *P*=1.9×10⁻⁵), and this association remained significant after adjustment for Aβ (β=0.085; 95% CI, 0.022 to 0.15; *P*=8.2×10⁻³; partial R^2^=0.0084) (**Figure 4A-C**). Similarly, GFAP mRNA expression was associated with longitudinal global cognitive slope (*n*=789, β=−0.13; 95% CI, −0.20 to −0.065; *P*=1.6×10⁻^4^), and this association persisted after adjustment for Aβ (β=−0.10; 95% CI, −0.17 to −0.034; *P*=3.3×10⁻³; partial R^2^=0.011). This association was no longer significant after additional adjustment for tau IHC (n=787, β=−0.059; 95% CI, −0.12 to 0.0013; *P*=0.055; partial R^2^=0.0047) (**Figure 4D-F**).

**Figure 4.**
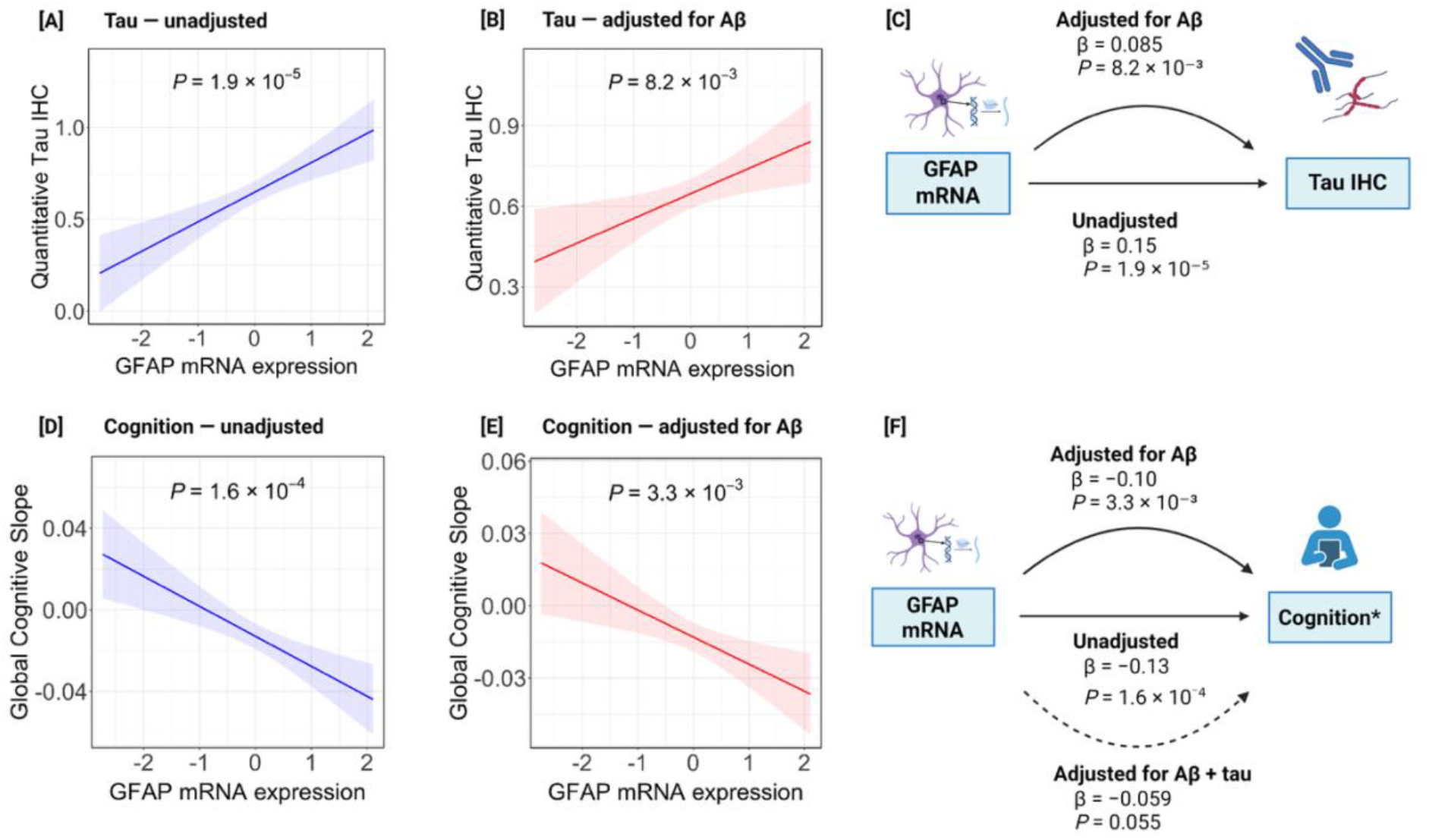
Association of brain GFAP mRNA expression with tau pathology and cognitive decline in ROSMAP. Linear regression models tested whether dorsolateral prefrontal cortex GFAP mRNA expression was associated with quantitative tau (immunohistochemistry [IHC]) and longitudinal global cognitive decline, before and after adjustment for amyloid-β (Aβ); GFAP mRNA expression was pre-residualized as described in Methods. In the plots, lines represent the fitted linear regression, shaded bands are 95% confidence intervals, and the annotated P value is for GFAP mRNA. In the diagrams, straight arrows denote the unadjusted association and curved arrows the adjusted associations; solid arrows indicate statistical significance (P<0.05) and dashed arrows nonsignificance. All β are standardized. [A] GFAP mRNA and tau IHC, unadjusted. [B] GFAP mRNA and tau IHC, adjusted for Aβ. [C] Association of GFAP mRNA with tau IHC before and after Aβ adjustment. [D] GFAP mRNA and global cognitive slope, unadjusted. [E] GFAP mRNA and global cognitive slope, adjusted for Aβ. [F] Association of GFAP mRNA with cognitive decline, unadjusted, after adjustment for Aβ, and after adjustment for both Aβ and tau IHC. **Abbreviations:** Aβ, amyloid-β; GFAP, glial fibrillary acidic protein; IHC, immunohistochemistry; mRNA, messenger RNA; ROSMAP, Religious Orders Study and Rush Memory and Aging Project. Created in BioRender. Danziger, R. (2026) https://BioRender.com/y6tpf65

To determine whether the GFAP-tau and GFAP-cognition associations varied by disease stage, we stratified by clinical diagnosis. GFAP mRNA was not associated with tau in participants who were cognitively unimpaired (*n*=270, *P*=0.64; after Aβ adjustment: *P*=0.90) or had MCI (*n*=211, *P*=0.80; after Aβ adjustment: *P*=0.69); we note both GFAP mRNA and tau burden were measured from the midfrontal cortex, which is rarely affected by tau pathology in CU individuals.^50,51^ Among participants with dementia (*n*=352), GFAP mRNA was associated with tau (β=0.27; 95% CI, 0.12 to 0.42; *P*=3.7×10⁻⁴), and this association persisted after adjustment for Aβ (β=0.19; 95% CI, 0.047 to 0.33; *P*=9.0×10⁻³) (**Supplementary Table 1**).

Similarly, GFAP mRNA was not associated with cognitive decline in those who were cognitively unimpaired (n=251, *P*=0.86; after Aβ adjustment: *P*=0.95) or had MCI (n=203, *P*=0.21; after Aβ adjustment: *P*=0.33). Among participants with dementia (n=335), GFAP mRNA was associated with faster cognitive decline (β=−0.13; 95% CI, −0.26 to −0.012; *P*=0.031). This association was moderately attenuated after adjustment for Aβ (β=−0.11; 95% CI, −0.24 to 0.0084; *P*=0.068) and further attenuated after adjustment for both Aβ and tau (β=−0.047; 95% CI, −0.16 to 0.064; *P*=0.41) (**Supplementary Table 2**).

### 3.5 Sensitivity analysis using silver-stain neuropathology

*APOE* ε4 was robustly associated with neuritic plaque burden (β=0.59; 95% CI, 0.44 to 0.74; *P*=4.2×10⁻¹⁴; F=59.1). *APOE* ε4 was also associated with GFAP mRNA expression (β=0.23; 95% CI, 0.080 to 0.39; *P*=2.9×10⁻³), but this association was no longer significant after adjustment for neuritic plaque (β=0.15; 95% CI, −0.010 to 0.31; P=0.067). TSLS analysis estimated the causal effect of neuritic plaque Aβ on GFAP mRNA expression (β=0.40; 95% CI, 0.13 to 0.67; P=3.5×10⁻³) (**Figure 5A**). GFAP mRNA expression was associated with NFT burden (β=0.15; 95% CI, 0.084 to 0.22; *P*=1.1×10⁻⁵), and this association remained significant after adjustment for neuritic plaque (β=0.080; 95% CI, 0.019 to 0.14; *P*=0.011). GFAP mRNA expression was associated with faster cognitive decline (β=−0.13; 95% CI, −0.20 to −0.065; *P*=1.6×10⁻⁴), and this association remained significant after adjustment for neuritic plaque (β=−0.088; 95% CI, −0.16 to −0.021; *P*=9.8×10⁻³). This association was no longer significant after adjusting for both neuritic plaque and NFT (β=−0.055; 95% CI, −0.12 to 0.0073; *P*=0.084) (**Supplementary Figure 2**).

**Figure 5.**
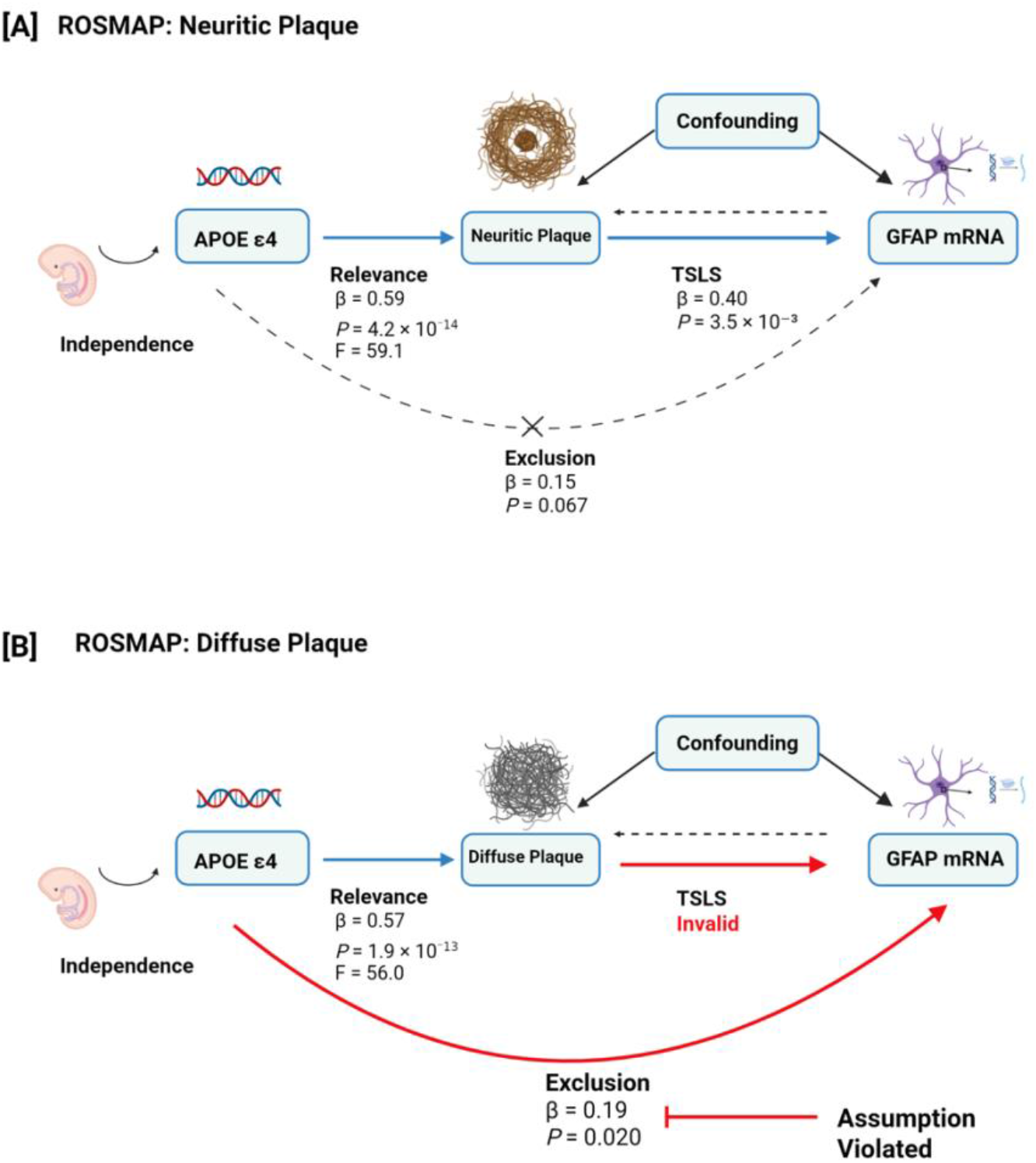
Mendelian randomization using silver-stain neuropathology: neuritic versus diffuse plaque (ROSMAP). Using APOE ε4 as the genetic instrumental variable, the causal effect of amyloid-β (Aβ) on GFAP mRNA expression was re-estimated with plaque burden quantified by modified Bielschowsky silver stain rather than immunohistochemistry. GFAP mRNA expression was pre-residualized as described in Methods. In the diagrams, solid arrows denote the instrument-exposure association (Relevance) and the two-stage least squares (TSLS) causal estimate; the backward dashed arrow denotes reverse causation (excluded by design); the crossed dashed arrow (X) is the exclusion-restriction path, for which the values shown are the APOE ε4-GFAP association after adjustment for plaque. The Confounding box denotes unmeasured confounders of the plaque-GFAP relationship, which the instrument is assumed to be independent of. Independence is supported by APOE ε4 being germline and fixed at conception. [A] Neuritic plaque: the APOE ε4-GFAP association was attenuated to nonsignificance after adjustment for neuritic plaque, consistent with the exclusion restriction, yielding a valid TSLS estimate. [B] Diffuse plaque: the APOE ε4-GFAP association remained significant after adjustment for diffuse plaque (curved red path), indicating that the exclusion restriction was violated; a valid causal estimate could not be obtained (red arrow). Together, these analyses support a causal effect of Aβ on GFAP for neuritic but not diffuse plaque. All β are standardized. **Abbreviations**: Aβ, amyloid-β; APOE, apolipoprotein E; GFAP, glial fibrillary acidic protein; mRNA, messenger RNA; ROSMAP, Religious Orders Study and Rush Memory and Aging Project; TSLS, two-stage least squares. Created in BioRender. Danziger, R. (2026) https://BioRender.com/fmgqita

*APOE* ε4 was robustly associated with diffuse plaque burden (β=0.57; 95% CI, 0.42 to 0.72; *P*=1.9×10⁻¹³; *F*=56.0). In contrast to the immunohistochemical and neuritic plaque analyses, the association of *APOE* ε4 with GFAP mRNA remained significant after adjustment for diffuse plaque (β=0.19; 95% CI, 0.031 to 0.35; *P*=0.020), indicating that the exclusion restriction was not satisfied. Because this assumption was violated, a valid causal estimate could not be obtained for diffuse plaque (**Figure 5B**). When neuritic and diffuse plaque were entered in the same model, GFAP mRNA expression was associated with neuritic plaque (β=0.17; 95% CI, 0.082 to 0.25; P=1.3×10⁻⁴) but not diffuse plaque (β=−0.0040; 95% CI, −0.089 to 0.081; P=0.93).

## 4.0 Discussion

Across two independent cohorts (HABS and ROSMAP), we integrated genetic, longitudinal in vivo, and postmortem evidence to clarify the Aβ-GFAP relationship. Using Mendelian randomization, we found consistent evidence in both cohorts supporting a causal effect of Aβ accumulation on GFAP elevation. This was true both for plasma GFAP in living, cognitively unimpaired adults and for neocortical GFAP expression in brain tissue at autopsy. The agreement between findings in plasma and brain tissue suggests that elevated GFAP reflects increased astrocytic GFAP production downstream of Aβ accumulation, rather than nonspecific release due to disease-related blood-brain barrier permeability or tissue injury. In histopathology, this causal effect was supported specifically when Aβ was measured as neuritic plaque but not diffuse plaque, suggesting that the GFAP elevation reflects an astrocytic response to the peri-plaque inflammation characteristic of neuritic amyloid.

A link between Aβ pathology and astrocytic GFAP is well recognized, though the evidence has been largely observational. In transgenic mouse models, amyloid deposition is accompanied by plaque-associated reactive astrocytes with increased GFAP expression.^52^ In human postmortem tissue, reactive astrocytes surround neuritic amyloid plaques, express GFAP,^53^ extend hypertrophic processes toward the plaque,^54^ and cortical GFAP expression rises with amyloid burden.^55^ Longitudinal cohort studies further establish the close relationship between plasma GFAP elevation and Aβ accumulation.^56,57^ Whereas these observations are associational, anti-amyloid therapy trials show that removing Aβ lowers plasma GFAP, which provides causal evidence in humans. Donanemab lowered plasma GFAP by 12%, with reductions proportional to plaque clearance.^23^ Gantenerumab, targeting fibrillar amyloid plaques, lowered plasma GFAP, whereas solanezumab, which targets soluble amyloid, did not reduce plaque burden or lower plasma GFAP.^58^ Our study builds on this prior work by providing human genetic evidence that Aβ drives increased GFAP transcription in brain tissue, specifically in relation to neuritic plaque, and, as a likely downstream consequence, increased plasma GFAP.

We then examined GFAP’s relationship with tau pathology and neurodegeneration. In HABS, baseline plasma GFAP was associated with longitudinal tau accumulation, but this relationship became nonsignificant after adjusting for Aβ. This indicates that in cognitively unimpaired individuals, plasma GFAP mainly reflects an astrocytic response to Aβ rather than tau. This is consistent with evidence that plasma GFAP is an early marker of amyloid but not tau pathology in the preclinical and early stages of disease.^11,56^ In ROSMAP brain tissue, by contrast, GFAP mRNA remained associated with tau pathology even after adjustment for Aβ, consistent with postmortem evidence in which serum GFAP was associated with tau pathology independently of amyloid,^20^ and this association was driven by the dementia stage. This suggests that at later disease stages, as tau burden rises, tau may make an additional contribution to astrocytic activation and GFAP expression. Plasma GFAP, measured at the preclinical stage, instead captures an earlier Aβ-dependent component of the astrocytic response that predominates before substantial tau burden accumulates. Finally, baseline plasma GFAP was not associated with longitudinal hippocampal atrophy, a key marker of neurodegeneration,^59,60^ either before or after adjustment for Aβ, in line with the inconsistent relationship between plasma GFAP and progressive neurodegeneration reported previously.^11,61^

The relationship between GFAP and cognition is more complex, as cognition is shaped by additional factors including vascular health, neuronal reserve, and environmental effects.^29,62^ In prior cohort studies, plasma GFAP predicted cognitive decline in CU adults who were amyloid positive,^61,63^ with little association in those who were amyloid negative.^61^ In HABS, plasma GFAP was associated with faster cognitive decline, but this was no longer significant after adjustment for Aβ, suggesting that its independent contribution to cognition in preclinical disease is modest and largely confounded by amyloid.^63,64^ In ROSMAP brain tissue, a prior study has shown that cortical GFAP expression is linked to faster cognitive decline.^55^ In our study, GFAP mRNA remained associated with cognitive decline even after adjustment for Aβ. However, this residual association lost significance after additional adjustment for tau, an effect that was most pronounced at the dementia stage. While reactive astrogliosis and downstream neuroinflammatory processes have been implicated in cognitive decline in AD,^8,22,64^ our findings from both cohorts indicate that GFAP itself largely reflects the burden of core AD pathology and carries little independent association with cognition once that pathology is accounted for. In ROSMAP, GFAP explained less than 1% of the remaining variance in cognitive decline after adjustment for Aβ and tau.

These findings align with the amyloid cascade model, in which aggregated Aβ drives tau phosphorylation and, as disease progresses, the combination of Aβ and tau in turn drives synaptic dysfunction, neurodegeneration, and glial activation, including reactive astrogliosis captured by elevated GFAP.^8,10^ Given our results, in preclinical disease plasma GFAP alone likely conveys little prognostic information beyond existing Aβ-associated markers such as p-tau217,^63^ although it might still have value in monitoring disease-modifying treatment responses should this become a standard of care in the future.^23,58^ GFAP is also a single marker and may not capture the diversity of reactive astrocytic states.^53^ Molecular and cellular profiling, including proteins downstream of astrocytic activation, is needed to determine whether markers more specific than GFAP alone can capture information about disease-associated neuroinflammation.

A major strength of this study is the replication of the MR findings across two independent cohorts that differ in design, disease stage, and measurement modality: in vivo amyloid PET and plasma GFAP in cognitively unimpaired adults, and postmortem neuropathology and brain GFAP expression across the clinical spectrum. The similar causal estimates obtained from these orthogonal measures make it unlikely that the findings reflect an artifact of any single assay or cohort. Our study has several limitations. Plasma GFAP is a nonspecific marker of astrocytic activation, and GFAP mRNA derived from bulk RNA sequencing reflects a composite signal across cell types.^17,45^ Both cohorts are predominantly white, and HABS enrolled only cognitively unimpaired adults, limiting generalizability to diverse populations or later disease stages, particularly as *APOE* ε4 confers stronger AD risk in white populations.^65^ To address this, a sensitivity analysis in ROSMAP restricted to participants of European ancestry yielded consistent MR results. In ROSMAP, GFAP expression and pathology were measured only in the DLPFC, a region affected by tau late in the disease course,^50^ hence the absence of an association between GFAP and tau in CU and MCI participants may reflect low regional tau burden. These findings also need to be replicated and clarified in earlier-involved limbic regions. Furthermore, causal inference itself relies on *APOE* ε4 as a single instrumental variable, limiting precision. In addition, Aβ burden was measured at a single time point in each cohort, so our estimates reflect the cumulative effect of genetically elevated Aβ and cannot distinguish whether the rate of deposition, impaired clearance, or duration of Aβ exposure is most relevant to GFAP elevation. *APOE* ε4 is also a pleiotropic genetic variant influencing neuroinflammation, lipid metabolism, and microglial function through pathways partially independent of Aβ, which could bias causal estimates.^66,67^ However, in both cohorts, the residual *APOE* ε4-GFAP association was no longer significant after accounting for Aβ, supporting the view that, in AD, most of the *APOE* ε4-GFAP link might be mediated through Aβ. Accordingly, our findings should be interpreted as genetic evidence consistent with causality rather than definitive mechanistic proof.^66,67^

In conclusion, across two independent cohorts, Mendelian randomization provides genetic evidence supporting a causal effect of Aβ accumulation on brain tissue GFAP expression and plasma GFAP elevation, likely reflecting an astrocytic response to neuritic plaque. Our results support the view that GFAP offers limited prognostic value beyond existing Aβ markers in preclinical AD. In postmortem tissue, GFAP was not independently associated with cognitive decline once Aβ and tau were accounted for. Further studies combining postmortem tissue evaluation across brain regions, in vivo biomarkers, and carefully designed mechanistic investigation are required to determine whether more specific astrocytic markers capture distinct disease-associated neuroinflammatory processes beyond core AD pathology.

## Contributors

RD: Design of the work; Analysis and interpretation of the data; Drafting the manuscript; Reviewing the work critically for important intellectual content

PK: Acquisition and interpretation of the data; Reviewing the work critically for important intellectual content

LT: Analysis and interpretation of the data; Reviewing the work critically for important intellectual content

ZY: Analysis and interpretation of the data; Reviewing the work critically for important intellectual content

JY: Analysis and interpretation of the data; Reviewing the work critically for important intellectual content

LY: Analysis and interpretation of the data; Reviewing the work critically for important intellectual content

JW: Analysis and interpretation of the data; Reviewing the work critically for important intellectual content

DHA: Analysis and interpretation of the data; Reviewing the work critically for important intellectual content

BJG-M: Analysis and interpretation of the data; Reviewing the work critically for important intellectual content

DVN: Analysis and interpretation of the data; Reviewing the work critically for important intellectual content

WWY: Analysis and interpretation of the data; Reviewing the work critically for important intellectual content

DJS: Acquisition and interpretation of the data; Reviewing the work critically for important intellectual content

JAS: Acquisition and interpretation of the data; Reviewing the work critically for important intellectual content

DAB: Acquisition and interpretation of the data; Reviewing the work critically for important intellectual content

PLD: Acquisition and interpretation of the data; Reviewing the work critically for important intellectual content

RFB: Acquisition and interpretation of the data; Reviewing the work critically for important intellectual content

KAJ: Acquisition and interpretation of the data; Reviewing the work critically for important intellectual content

RAS: Acquisition and interpretation of the data; Reviewing the work critically for important intellectual content

SEA: Acquisition and interpretation of the data; Reviewing the work critically for important intellectual content

JPC: Conception and design of the work; Acquisition, analysis, and interpretation of the data; Reviewing the work critically for important intellectual content

H-SY: Conception and design of the work; Acquisition, analysis, and interpretation of the data; Drafting the work; Reviewing the work critically for important intellectual content

## Data Availability

ROSMAP data, including all data used in this study (e.g., deidentified participant-level data, data dictionary), can be requested at the online RADC Research Resource Sharing Hub (https://www.radc.rush.edu). The requesting investigators are required to complete and submit a formal proposal as outlined in the online RADC Research Resource Sharing Hub. The data will be provided after the data request is reviewed by the RADC and the Data Use Agreement is signed.
Harvard Aging Brain Study (HABS) data, including deidentified participant-level data (demographic, clinical, neuropsychological, biomarker, and neuroimaging), are publicly available and can be requested through the HABS website (https://habs.mgh.harvard.edu/researchers/request-data/), with the current public data release distributed via Synapse (https://www.synapse.org/habs). Requesting investigators are required to agree to the Harvard Aging Brain Study Data Use Agreement. The data are provided after the request is reviewed and approved by the HABS data managers and data committee.

https://www.synapse.org/habs

## Acknowledgments

We thank the ROSMAP study participants and staff of the Rush Alzheimer’s Disease Center. We thank the Harvard Aging Brain Study participants and staff. The Harvard Aging Brain Study is conducted through the Department of Neurology at Massachusetts General Hospital and the Center for Alzheimer Research and Treatment at Brigham and Women’s Hospital, with neuroimaging performed at the Athinoula A. Martinos Center for Biomedical Imaging, Massachusetts General Hospital, Charlestown, Massachusetts. We thank Meso Scale Discovery for providing assay kits free of charge to measure AD biomarkers in HABS. Figures 2,3,4,5 and Supplementary Figure 2 were created with BioRender.com. Publication licenses are listed in each figure legend.

## Data Sharing

ROSMAP data, including all data used in this study (e.g., deidentified participant-level data, data dictionary), can be requested at the online RADC Research Resource Sharing Hub (https://www.radc.rush.edu). The requesting investigators are required to complete and submit a formal proposal as outlined in the online RADC Research Resource Sharing Hub. The data will be provided after the data request is reviewed by the RADC and the Data Use Agreement is signed.

Harvard Aging Brain Study (HABS) data, including deidentified participant-level data (demographic, clinical, neuropsychological, biomarker, and neuroimaging), are publicly available and can be requested through the HABS website (https://habs.mgh.harvard.edu/researchers/request-data/), with the current public data release distributed via Synapse (https://www.synapse.org/habs). Requesting investigators are required to agree to the Harvard Aging Brain Study Data Use Agreement. The data are provided after the request is reviewed and approved by the HABS data managers and data committee.

## SUPPLEMENTARY TABLES AND FIGURES

**Supplementary Table 1.** Stage-stratified association between GFAP mRNA expression and tau pathology (midfrontal tau immunohistochemistry), before and after adjustment for amyloid-β.

| **Clinical stage** | ***n*** | **Unadjusted** |  | **Aβ-adjusted** |  |
|---|---|---|---|---|---|
|  |  | **β (95% CI)** | ***P*** | **β (95% CI)** | ***P*** |
| All participants | 833 | 0.15 (0.080 to 0.21) | 1.9 × 10<sup>-5</sup> | 0.085 (0.022 to 0.15) | 8.2 × 10<sup>-3</sup> |
| Cognitively unimpaired | 270 | 0.010 (−0.032 to 0.052) | 0.64 | −0.0027 (−0.043 to 0.038) | 0.90 |
| MCI | 211 | 0.012 (−0.083 to 0.11) | 0.80 | −0.018 (−0.11 to 0.069) | 0.69 |
| Dementia | 352 | 0.27 (0.12 to 0.42) | 3.7 × 10<sup>-4</sup> | 0.19 (0.047 to 0.33) | 9.0 × 10<sup>-3</sup> |

β is the standardized GFAP mRNA coefficient with 95% CI from linear models of midfrontal quantitative tau IHC (outcome) on GFAP mRNA expression (predictor), in all participants and within each clinical stage. Variables were standardized in the overall sample. DLPFC GFAP mRNA was pre-residualized as described in Methods. Clinical stage was defined by final consensus diagnosis.
**Abbreviations**: Aβ, amyloid-β; CI, confidence interval; DLPFC, dorsolateral prefrontal cortex; GFAP, glial fibrillary acidic protein; IHC, immunohistochemistry; MCI, mild cognitive impairment.

**Supplementary Table 2.** Stage-stratified association between GFAP mRNA expression and the rate of cognitive decline (global cognitive slope), unadjusted and after sequential adjustment for amyloid-β and tau.

| **Clinical stage** | ***n*** | **Unadjusted** |  | **Aβ-adjusted** |  | **Aβ + tau-adjusted** |  |
|---|---|---|---|---|---|---|---|
|  |  | **β (95% CI)** | ***P*** | **β (95% CI)** | ***P*** | **β (95% CI)** | ***P*** |
| All participants | 789 | −0.13 (−0.20 to −0.065) | 1.6 × 10<sup>-4</sup> | −0.10 (−0.17 to −0.034) | 3.3 × 10<sup>-3</sup> | −0.059 (−0.12 to 0.0013) | 0.055 |
| Cognitively unimpaired | 251 | 0.0044 (−0.044 to 0.052) | 0.86 | 0.0016 (−0.046 to 0.050) | 0.95 | 0.0029 (−0.045 to 0.051) | 0.91 |
| MCI | 203 | −0.043 (−0.11 to 0.025) | 0.21 | −0.034 (−0.10 to 0.034) | 0.33 | −0.038 (−0.10 to 0.028) | 0.26 |
| Dementia | 335 | −0.13 (−0.26 to −0.012) | 0.031 | −0.11 (−0.24 to 0.0084) | 0.068 | −0.047 (−0.16 to 0.064) | 0.41 |

β is the standardized GFAP mRNA coefficient with 95% CI from linear models of the global cognitive slope (outcome) on GFAP mRNA expression (predictor), in all participants and within each clinical stage. Variables were standardized in the overall sample. The Aβ + tau-adjusted column uses the subset with non-missing midfrontal tau IHC (all participants, n=787; cognitively unimpaired, n=250; MCI, n=202; dementia, n=335). DLPFC GFAP mRNA was pre-residualized as described in Methods. Clinical stage was defined by final consensus diagnosis.
**Abbreviations**: Aβ, amyloid-β; CI, confidence interval; DLPFC, dorsolateral prefrontal cortex; GFAP, glial fibrillary acidic protein; IHC, immunohistochemistry; MCI, mild cognitive impairment.

---

**Supplementary Figure 1.**
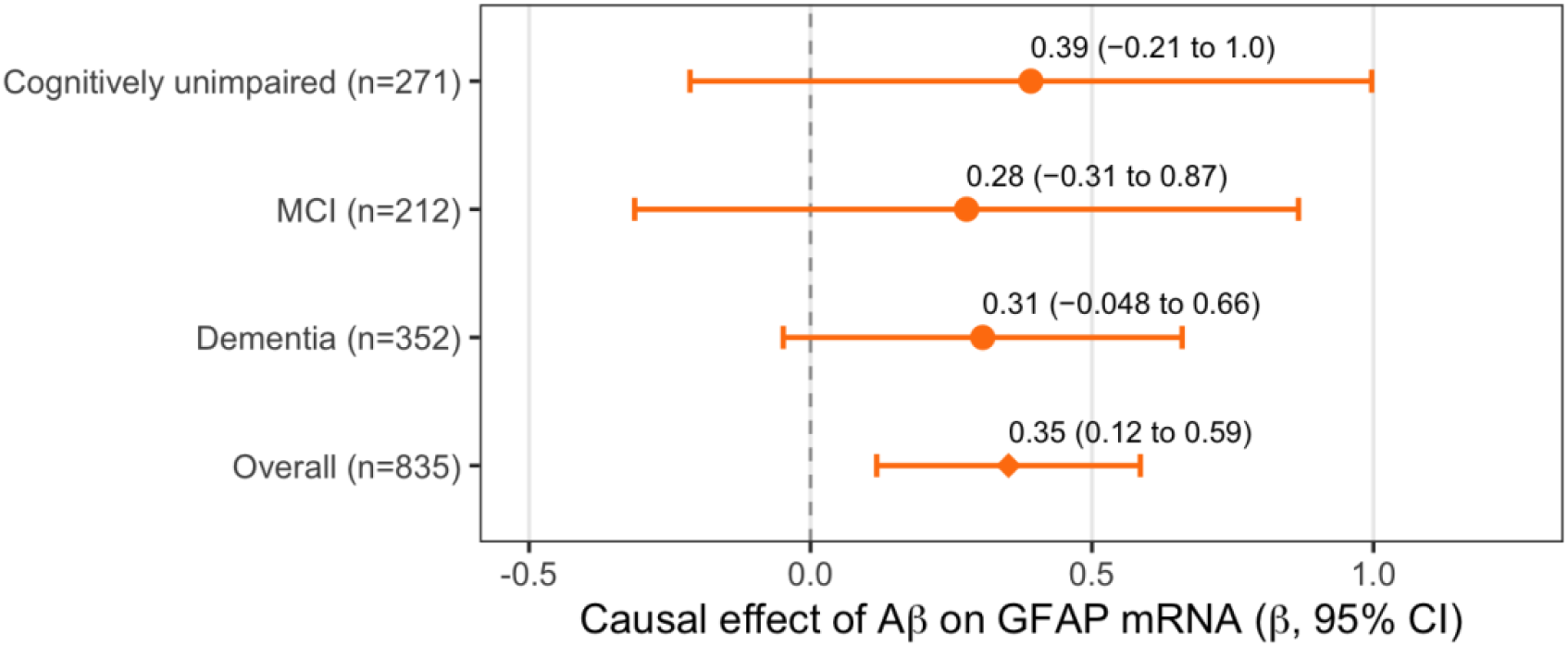
Mendelian randomization of amyloid-β on GFAP mRNA expression stratified by clinical diagnosis (ROSMAP). Two-stage least squares (TSLS) estimates of the causal effect of amyloid-β (Aβ) on dorsolateral prefrontal cortex GFAP mRNA expression, using APOE ε4 carrier status as the genetic instrumental variable and quantitative Aβ immunohistochemistry as the exposure. Estimates are shown separately for participants who were cognitively unimpaired, had mild cognitive impairment (MCI), or had dementia at the final clinical evaluation, and for the overall sample. Points are standardized β estimates and horizontal bars are 95% confidence intervals; circles denote stage-specific estimates, and the diamond denotes the overall estimate. The vertical dashed line indicates no effect. Aβ and GFAP mRNA were standardized in the overall sample, and GFAP mRNA expression was pre-residualized as described in Methods. First-stage F statistics were 16.1 (cognitively unimpaired), 10.7 (MCI), 30.5 (dementia), and 77.8 (overall). Point estimates were similar across stages and consistent with the overall estimate, although none reached statistical significance within an individual stage. **Abbreviations**: Aβ, amyloid-β; APOE, apolipoprotein E; CI, confidence interval; GFAP, glial fibrillary acidic protein; MCI, mild cognitive impairment; mRNA, messenger RNA; ROSMAP, Religious Orders Study and Rush Memory and Aging Project; TSLS, two-stage least squares.

**Supplementary Figure 2.**
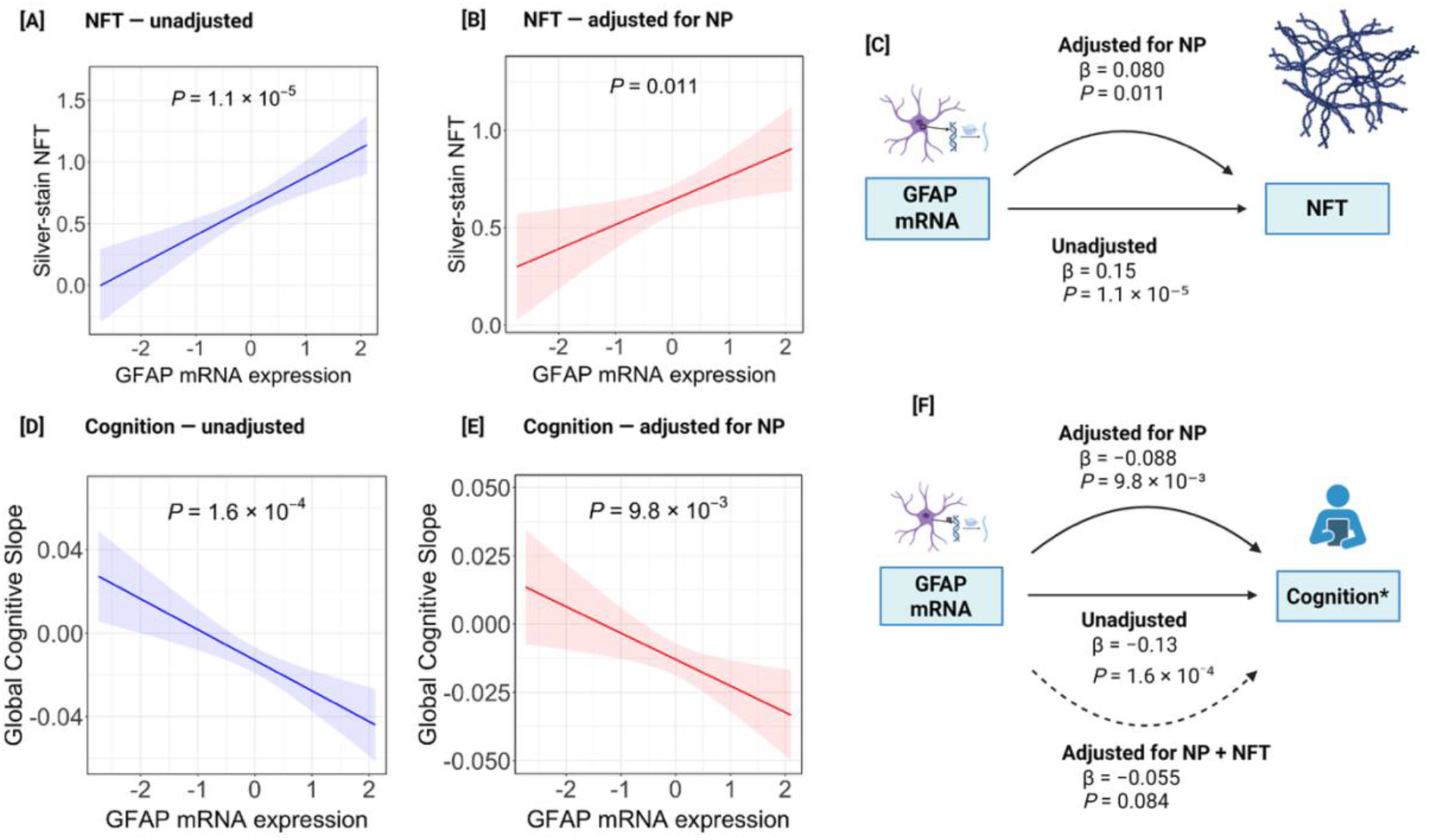
Association of brain GFAP mRNA expression with neurofibrillary tangles and cognitive decline using silver-stain neuropathology (ROSMAP). Linear regression models tested whether dorsolateral prefrontal cortex GFAP mRNA expression was associated with midfrontal neurofibrillary tangle (NFT) burden and longitudinal global cognitive decline, before and after adjustment for neuritic plaque (NP) burden. NFT and NP were counted on modified Bielschowsky silver stain and square-root transformed. GFAP mRNA expression was pre-residualized as described in Methods. In the plots, lines represent the fitted linear regression, shaded bands are 95% confidence intervals, and the annotated P value is for GFAP mRNA. In the diagrams, straight arrows denote the unadjusted association and curved arrows the adjusted associations; solid arrows indicate statistical significance (P<0.05) and dashed arrows nonsignificance. All β are standardized. [A] GFAP mRNA and NFT, unadjusted. [B] GFAP mRNA and NFT, adjusted for NP. [C] Association of GFAP mRNA with NFT before and after NP adjustment. [D] GFAP mRNA and global cognitive slope, unadjusted. [E] GFAP mRNA and global cognitive slope, adjusted for NP. [F] Association of GFAP mRNA with cognitive decline, unadjusted, after adjustment for NP, and after adjustment for both NP and NFT. **Abbreviations**: GFAP, glial fibrillary acidic protein; mRNA, messenger RNA; NFT, neurofibrillary tangle; NP, neuritic plaque; ROSMAP, Religious Orders Study and Rush Memory and Aging Project. Created in BioRender. Danziger, R. (2026) https://BioRender.com/1t7plzk

